## Supplementary Material for "Potential impact, costs, and benefits of population-wide screening interventions for tuberculosis in Viet Nam: a mathematical modelling study"

1. TB Modelling Group, TB Centre, London School of Hygiene and Tropical Medicine, London, United Kingdom; 2. Department of Infectious Disease Epidemiology, London School of Hygiene and Tropical Medicine, London, United Kingdom; 3. Instituto de Medicina Tropical Alexander von Humboldt, Universidad Peruana Cayetano Heredia, Lima, Peru; 4. Global Health Economics Centre, London School of Hygiene and Tropical Medicine, London, United Kingdom; 5. Department of Internal Medicine and Radboud Community for Infectious Diseases, Radboud University Medical Center, Nijmegen, the Netherlands; 6. South West Sydney Clinical Campuses, University of New South Wales, Sydney, Australia; 7. Ingham Institute of Applied Medical Research, Sydney, Australia; 8. Ministry of Health, Ha Noi, Viet Nam; 9. The University of Sydney Vietnam Institute, Ho Chi Minh City, Viet Nam; 10. Faculty of Medicine and Health, University of Sydney, Sydney, Australia; 11. The University of Sydney Institute for Infectious Diseases, Sydney, Australia; 12. Woolcock Institute of Medical Research, Sydney, Australia; 13. Department of Global Health, Amsterdam University Medical Centers, University of Amsterdam, Amsterdam, the Netherlands; 14. Amsterdam Institute for Global Health and Development, Amsterdam, the Netherlands; 15. National Lung Hospital, National Tuberculosis Control Programme, Ha Noi, Viet Nam; 16. School of Clinical Medicine, University of New South Wales, Sydney, Australia; 17. Burnet Institute, Melbourne, Australia.

**Keywords:** active detection; cost-effectiveness; subclinical; TB elimination

**Table of Contents:**

*Supplementary Methods:* ... *3*

SM1. Baseline model structure ... 3

SM2. Baseline model equations ... 4

SM3. Calibration methodology ... 5

SM4. Screening model structure ... 5

SM5. Disability-adjusted life years calculations ... 6

*Supplementary Figures:* ... *7*

Figure S1. TB natural history model under population-wide screening. ... 7

Figure S2. TB prevalence rate reduction under population-wide screening algorithms. ... 8

Figure S3. Proportional reduction of TB prevalence under NAAT-only approach and ACT3. ... 9

Figure S4. Incremental costs under CXR+NAAT algorithm. ... 10

*Supplementary Tables:* ... *11*

Table S1. Calibration targets. ... 11

Table S2. Model parameter description, ranges, and non-implausible points. ... 12

Table S3. Probability of a positive test per model state for each screening tool. ... 14

Table S4. Costing estimates for business-as-usual TB diagnosis and treatment. ... 16

Table S5. Costing estimates for population-wide screening algorithms. ... 17

Table S6. Performance of population-wide screening interventions to reach TB prevalence threshold of 100 per 100,000 inhabitants. ... 18

Table S7. Performance of population-wide screening interventions to reach TB prevalence threshold of 20 per 100,000 inhabitants. ... 19

Table S8. Performance of population-wide screening interventions with further investigation post-screening. ... 20

Table S9. Performance of population-wide screening interventions with revised CXR sensitivity. ... 21

Table S10. Cost-effectiveness of population-wide screening interventions for TB. ... 22

*References* ... *24*

**Supplementary Methods:**

**SM1. Baseline model structure**

We developed a compartmental model of tuberculosis (TB) natural history, adapting features of previously published models [1]. The model structure is shown in **Figure 1** on the manuscript. Model parameters used and their definitions are provided in **Table S2** in a later section within this document. This model was run from 1500 to 2020 with some time-varying parameters. The model tracked a closed population of 100,000 adults (≥15 years old).

We represented TB natural history with nine distinct compartments allowing for *Mtb* infection through an annual risk of infection (ARI). Disease state classification was informed by the ICE-TB framework, and naming follows current World Health Organization (WHO) definitions [2,3]:

- Unconfirmed TB disease (uTB): individuals with inflammatory pathology (evidenced through imaging methods) prior to the onset of bacteriological evidence of TB disease or symptoms of active TB disease.
- Asymptomatic TB disease (aTB): individuals with bacteriological evidence of TB disease who do not report symptoms of active TB disease on screening.
- Symptomatic TB disease (sTB): individuals with bacteriological evidence of TB disease with symptoms of active TB disease.
- Infectious disease: refers to bacteriologically positive disease; as such, it includes asymptomatic and symptomatic disease.
- TB disease: Any state of unconfirmed, asymptomatic, or symptomatic disease.

The ARI $\lambda$ depends upon the contact parameter $\beta$ and the prevalence of infectious disease (i.e., asymptomatic and symptomatic TB). Additionally, the relative infectiousness $\kappa$ of asymptomatic TB is also considered. The formula for the ARI $\lambda$ is presented later in this document. Individuals in the *Susceptible* (S) compartment could become infected with *Mtb* and progress to the *Infection* (I) compartment. From *Infection* (I), three pathways are possible: (i) self-clearance of infection (i.e. *Cleared* (C)) at rate *infcle*, (ii) progression to *Unconfirmed* (uTB) at rate *infunc*, and (iii) progression to *Asymptomatic* (aTB) at rate *infasy*. TB transmission in the model (i.e., transition into the *Infected* (I) compartment) can occur via the ARI $\lambda$ through four distinct routes: through first infection from *Susceptible* (S), through reinfection after self-clearance from *Cleared* (C), through reinfection after self-cure from *Recovered* (R) accounting for protection from reinfection $\pi$, and through reinfection after treatment from *Treated* (Tr) accounting for increased risk of reinfection $\rho$. TB disease states are sequentially depicted in the model in the *Unconfirmed* (uTB), *Asymptomatic* (aTB), and *Symptomatic* (sTB) compartments, allowing progression (denoted with parameters *uncasy* and *asysym*) and regression (denoted with parameters *asyunc* and *symasy*). For the *Unconfirmed* (uTB) compartment, individuals can transition out of disease states by self-cure into the *Recovered* (R) compartment at rate *uncrec*; here, reinfection can occur via the ARI but we assume there is protection from reinfection $\pi$. The model assumes that TB diagnosis and treatment only occur for individuals in the *Symptomatic* (sTB) compartment at rate $\theta$. Furthermore, it accounts for TB-specific mortality $\mu_{TB}$ in this compartment. Once in *Treatment* (Tx), there can be treatment failure at rate $\phi$ and treatment completion at rate $\delta$. Finally, in the *Treated* (Tr) compartment, reinfection can occur via the ARI $\lambda$ parameter.

The model also accounts for background mortality having a fixed rate $\mu$ (representing an age expectancy of 70 years) in each compartment. As a closed population model, the sum of all background and TB-specific mortality is fed back into the *Susceptible* compartment through the $\omega$ parameter.

**SM2. Baseline model equations**

A series of ordinary differential equations were set in place to represent the model structure mathematically. Parameter symbols and descriptions are outlined in **Table S2**. Parameter names indicate direction (i.e., *infcle* denotes from *Infection* to *Cleared*), and subscript *t* denotes parameters that vary over time. All nine compartments are represented: *Susceptible* (S), *Infected* (I), *Cleared* (C), *Recovered* (R), *Unconfirmed* (uTB), *Asymptomatic* (aTB), *Symptomatic* (sTB), *Treatment* (Tx), *Treated* (Tr). Given N = 100,000, then:

$$\frac{dS}{dt} = \mu \cdot (N - S) + \mu_{TB,t} \cdot sTB - \lambda \cdot S$$

$$\frac{dI}{dt} = \lambda \cdot (S + C + \pi \cdot R + \rho \cdot Tr) - I \cdot (infcle + infunc + infasy + \mu)$$

$$\frac{dC}{dt} = infcle \cdot I - C \cdot (\lambda + \mu)$$

$$\frac{dR}{dt} = uncrec \cdot uTB - R \cdot (\lambda \cdot \pi + \mu)$$

$$\frac{duTB}{dt} = infunc \cdot I + asyunc \cdot aTB - uTB \cdot (uncrec + uncasy + \mu)$$

$$\frac{daTB}{dt} = I \cdot (infasy + uncasy) + symasy \cdot sTB - aTB \cdot (asyunc + asysym + \mu)$$

$$\frac{dsTB}{dt} = asysym \cdot aTB - sTB \cdot \left(symasy + \theta_t + \mu_{TB,t} + \mu\right) + \varphi_t \cdot Tx$$

$$\frac{dTx}{dt} = \theta_t \cdot sTB - Tx \cdot (\varphi_t + \delta + \mu)$$

$$\frac{dTr}{dt} = \delta \cdot Tx - Tr \cdot (\lambda \cdot \rho + \mu)$$

As mentioned before, the ARI $\lambda$ depends upon the contact parameter $\beta$ and the prevalence of infectious disease (i.e., asymptomatic and symptomatic TB) and its equation is expressed below.

$$\lambda = \frac{\beta \cdot (\kappa \cdot aTB + sTB)}{N}$$

**SM3. Calibration methodology**

We calibrated the model using history matching with emulation, a calibration method that explores high-dimensional parameter spaces efficiently [4]. History matching refers to the exploration of the ranges of parameters given and identifying parameter sets that give rise to model outputs that match empirical data [4]. History matching progresses through multiple iterations (referred to as waves), where implausible areas of parameters (i.e., values where no match is found) are identified and discarded [4]. This process is made efficient with the use of emulators, which provide approximations of model outputs orders of magnitudes faster than the model [4]. As a result of multiple waves, the implausible space is reduced, resulting in parameter sets that match calibration targets.

History matching with emulation was implemented using the *hmer* package in R [5]. Calibration targets were TB epidemiological and demographic data of Viet Nam (S1 Table). The model comprised 23 dynamic parameters which are described in **Table S2**. The parameter ranges (priors) and sources are outlined. The non-implausible points (posteriors) were calculated as the median and corresponding 95% uncertainty intervals, calculated as the 2.5th to 97.5th percentiles of the parameter sets.

**SM4. Screening model structure**

We expanded the baseline TB natural history model described above to incorporate population-wide screening interventions from 2020 to 2050. The model structure remains mostly unchanged except for the transitions which occur during the years where the screening is applied in various annual rounds from 2025 (see dashed lines in **Figure S2**). When screened, each compartment transitions into a treatment compartment and after completion transitions back into its original compartment except *Infection* (I) which transitions into *Cleared* (C) and the disease compartments (*Unconfirmed* (uTB), *Asymptomatic* (aTB), and *Symptomatic* (sTB)) which transition into *Treated* (Tr). Individuals in the *Treatment* (Tx) compartment are not screened as part of screening interventions. The rates of transitions per compartment are outlined in **Table 1** according to the tool used. The model is no longer a closed-population model and now accounts for birth and mortality rates for Viet Nam from 2020 to 2050 [6].

**SM5. Disability-adjusted life years calculations**

Since our compartmental model does not track ageing, we opted to estimate mean lifetime disability-adjusted life years (DALYs) per incident TB. For this, we used the total DALYs for TB disease (0.40 million; 95%CI: 0.33-0.49) and post-TB (0.85 million; 95%CI: 0.57-1.22) in Viet Nam in 2019, as estimated by Menzies et al [6]. Then, considering the number of incident TB estimated in Viet Nam in 2019 (169,000), we calculated the point value for lifetime DALYs per incident TB: 2.4 (95%CI: 2.0-2.9) for TB disease and 5.0 (95%CI: 3.4-7.2) for post-TB [7,8].

To estimate the DALYs lived with post-TB, we used the weighted average age of individuals with TB in Viet Nam (49 years) from the WHO Global TB Report and obtained the life expectancy at that age (29.5 years) from the United Nations World Population Prospects [6,8]. Next, we estimated the proportion of an individual's remaining lifetime that would occur between the start of the population-wide screening interventions in 2025 and the time horizon of 2050, a period chosen to align with the duration of the implementation of the intervention and evaluation timeframe. Ultimately, lifetime DALYs per incident TB were calculated as the DALYs due to TB disease episode plus the DALYs lived with post-TB, discounted at a rate of 3% per year from 2025 [9].

**Supplementary Figures:**

**Figure S1. TB natural history model under population-wide screening.**

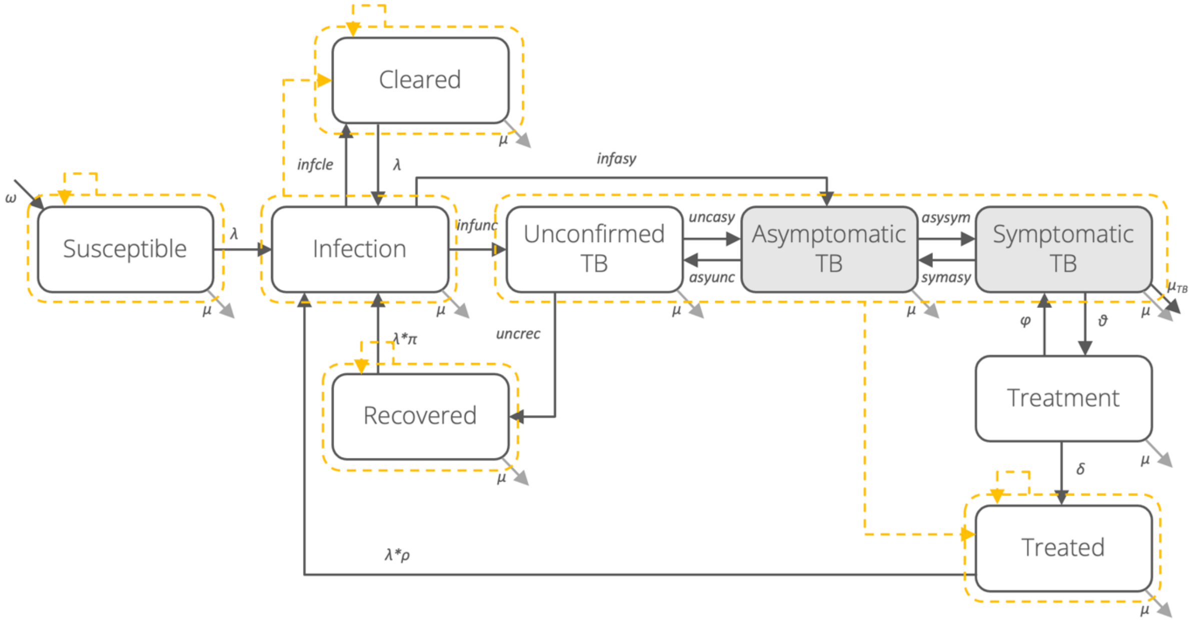

A compartmental model of tuberculosis natural history, adapting features of previously published models [1,10]. The model is depicted using nine compartments allowing for *Mycobacterium tuberculosis* infection through. Shaded compartments indicate those that contribute to transmission. The force of infection (depicted with λ) depends upon the contact parameter and the prevalence of infectious disease (i.e., asymptomatic and symptomatic TB), accounting for the relative infectiousness of asymptomatic TB. Dashed lines symbolise compartment flow after TB treatment.

**Figure S2. TB prevalence reduction under population-wide screening algorithms.**

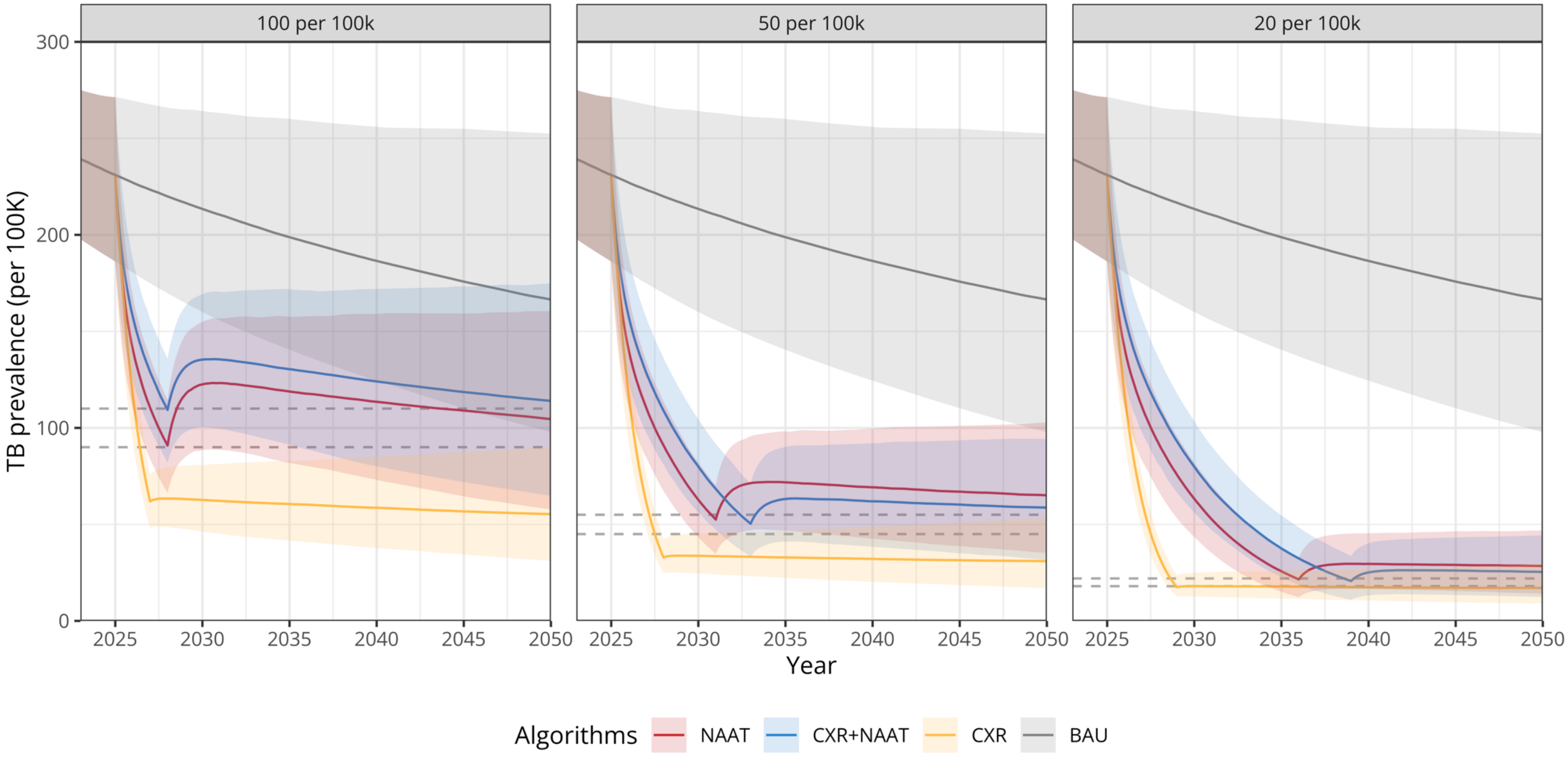

TB prevalence by each population-wide screening algorithm per threshold. Main analysis evaluates performance to reach TB prevalence of 50 per 100,000 people. Lines represent TB prevalence, and the shaded area shows the lower (2.5% quantile) and upper (97.5% quantile) bounds. Dashed lines represent TB prevalence thresholds (±10%). NAAT: Nucleic acid amplification test (Xpert MTB/RIF Ultra); CXR: Chest radiography with CAD software interpretation.

**Figure S3. Proportional reduction of TB prevalence under NAAT-only approach and ACT3.**

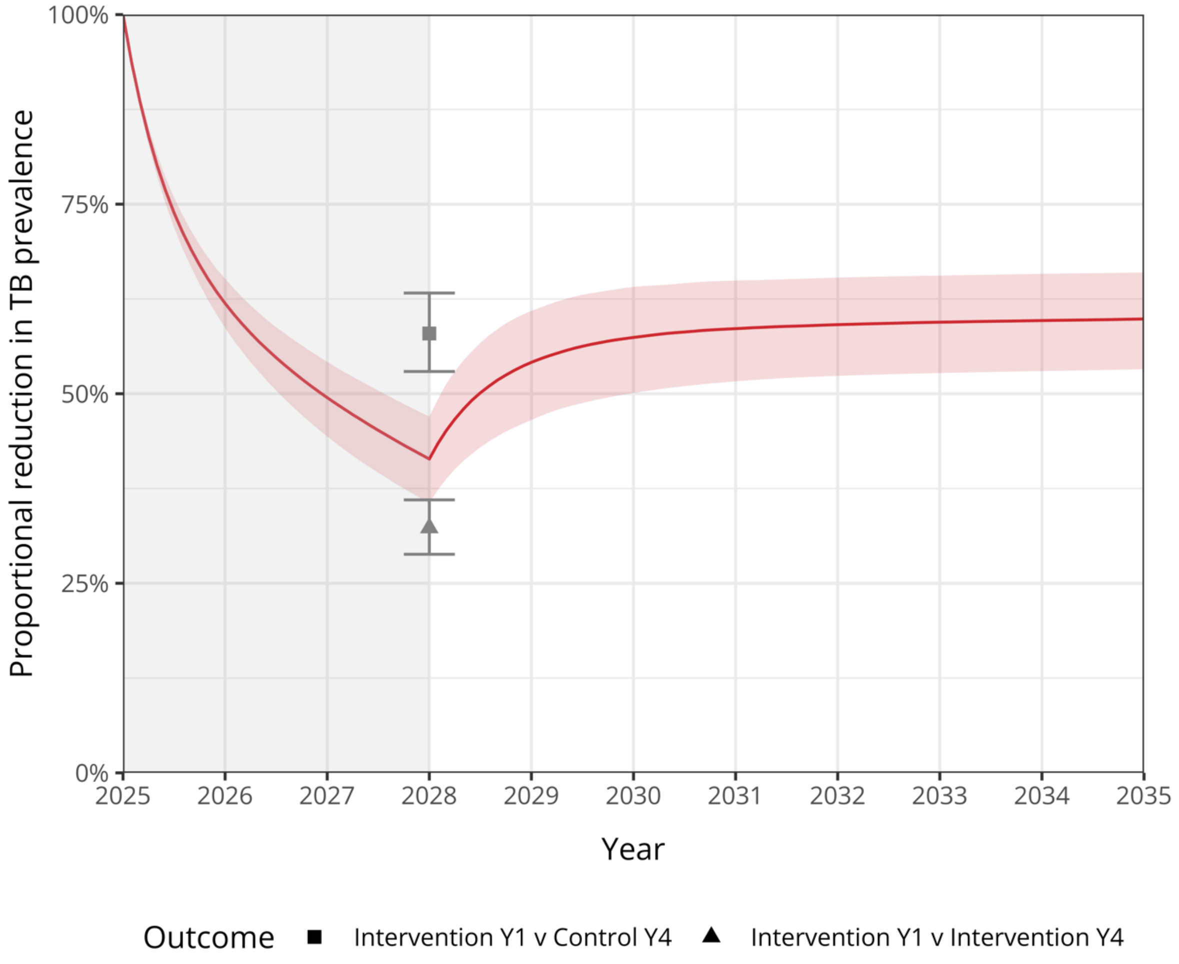

Proportional TB prevalence reduction under three annual rounds of NAAT-only algorithm compared to business-as-usual. Line represents proportional TB prevalence reduction per 100,000 people, and the shaded area shows the lower (2.5% quantile) and upper (97.5% quantile) bounds. The symbols and error bars represents TB prevalence proportional reduction as observed in the ACT3 trial [11].

**Figure S4. Incremental costs under CXR+NAAT algorithm.**

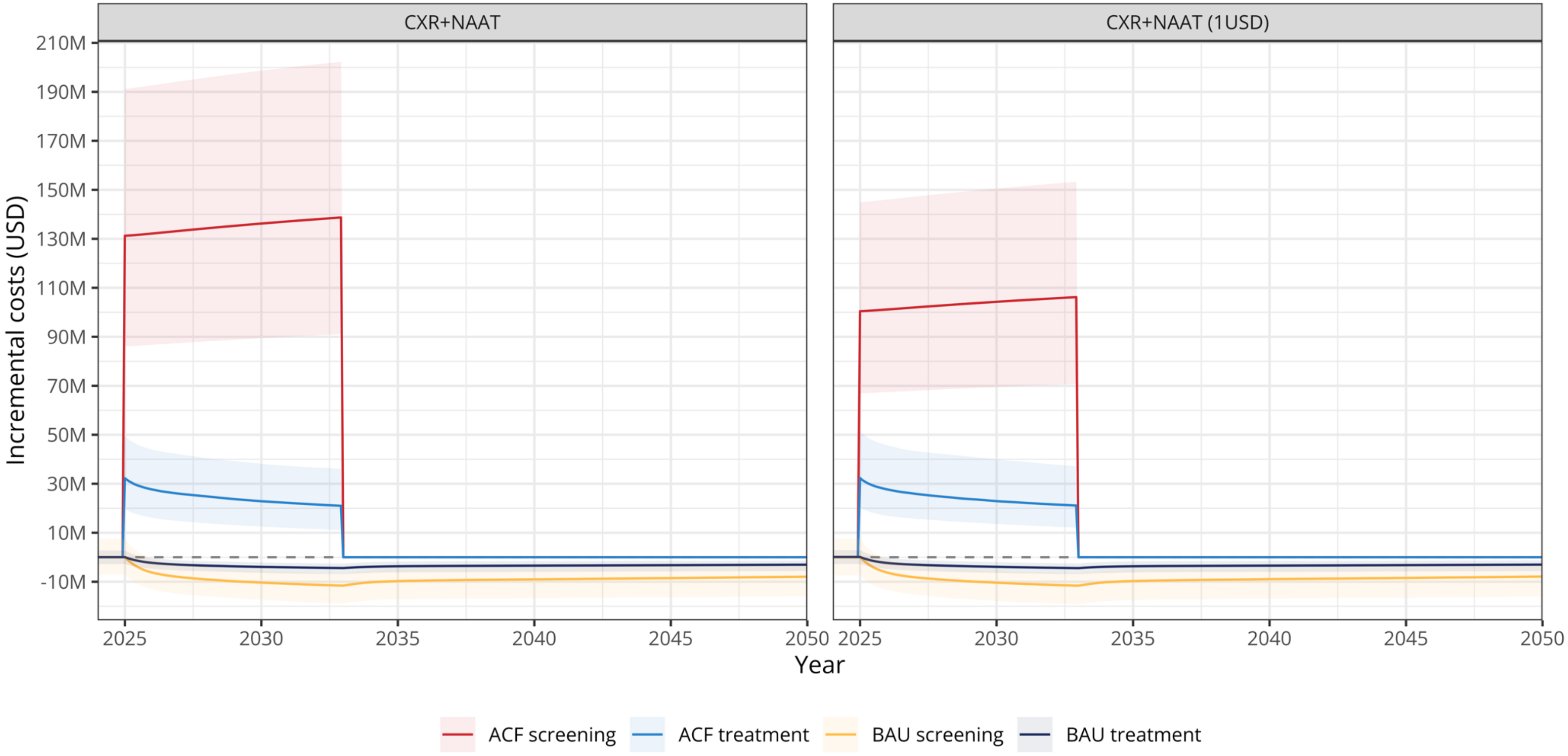

Incremental costs compared to the business-as-usual (BAU) counterfactual of population-wide screening interventions in Viet Nam using a CXR+NAAT algorithm to achieve a TB prevalence of 50 per 100,000 people. Incremental costs are shown by category, with ACF representing intervention-specific costs and BAU reflecting business-as-usual TB prevention and care costs. The main analysis assumed an Xpert MTB/RIF Ultra cartridge cost of 8 USD, while a sensitivity analysis explored a reduced cartridge cost of 1 USD. Lines represent incremental costs, and the shaded areas indicate the lower (2.5% quantile) and upper (97.5% quantile) bounds.

**Supplementary Tables:**

**Table S1. Calibration targets.**

| Target | Year | Value [95%CI] | Source |
|---|---|---|---|
| TB prevalence per 100,000 people | 2007 | 250 [202 – 310] | [12] |
| | 2018 | 227 [177 – 290] | [12] |
| TB mortality rate per 100,000 people | 2000 | 59.7 [37.3 - 87.7] | [6,8] |
| | 2010 | 33.5 [22.8 - 45.7] | [6,8] |
| TB notification rate per 100,000 people | 2010 | 79.4 [63.5 - 95.3] | [6,8] |
| | 2020 | 73.2 [58.5 - 87.8] | [6,8] |
| Proportion asymptomatic TB | 2007 | 0.70 [0.56 - 0.84] | [13] |
| | 2018 | 0.66 [0.53 - 0.79] | [13] |

Calibration targets, with brackets indicating 95% confidence intervals (95%CI), for TB epidemiology in Viet Nam were set for the adult population aged 15 years and older. TB prevalence refers specifically to infectious TB (i.e., asymptomatic and symptomatic), TB mortality is specific to deaths from symptomatic TB, TB notification represents the number of individuals with symptomatic TB initiating treatment through the business-as-usual approach, and the proportion of asymptomatic TB refers to the proportion of all infectious TB that is asymptomatic. Estimates for the TB prevalence used as calibration targets differ from those available in reference; since publication, an observed disparity in the proportional decline was corrected by the authors, and the estimates provided here reflect this correction.

**Table S2. Model parameter description, ranges, and non-implausible points.**

| Parameters | Description | Ranges | Non-implausible ranges [95%UI] | Sources |
|---|---|---|---|---|
| beta (β) | Transmission coefficient | 6.00 - 20.00 | 14.16 [8.82 - 19.29] | - |
| kappa (κ) | Relative transmission from asymptomatic TB | 0.62 - 1.00 | 0.82 [0.64 - 0.98] | [13] |
| pi (π) | Relative risk of reinfection after recovery from unconfirmed TB | 0.14 - 0.30 | 0.21 [0.15 - 0.29] | [14] |
| rho (ρ) | Relative risk of reinfection after treatment completion | 2.14 - 4.27 | 3.15 [2.23 - 4.19] | [15] |
| infcle | Rate of clearance from infection per year | 0.93 - 3.30 | 1.90 [1.09 - 2.94] | [1] |
| infunc | Rate of progression from infection to unconfirmed TB per year | 0.04 - 0.23 | 0.16 [0.06 - 0.22] | [1] |
| infasy | Rate of progression from infection to asymptomatic TB per year | 0.01 - 0.10 | 0.06 [0.01 - 0.10] | [1] |
| uncrec | Rate of recovery from unconfirmed TB per year | 0.14 - 0.23 | 0.18 [0.14 - 0.22] | [1] |
| uncasy | Rate of progression from unconfirmed to asymptomatic TB per year | 0.21 - 0.28 | 0.25 [0.21 - 0.28] | [1] |
| asyunc | Rate of recovery from asymptomatic to unconfirmed TB per year | 1.24 - 2.03 | 1.66 [1.30 - 1.99] | [1] |
| asysym | Rate of progression from asymptomatic to symptomatic TB per year | 0.56 - 0.94 | 0.88 [0.76 - 0.94] | [1] |
| symasy | Rate of recovery from symptomatic to asymptomatic TB per year | 0.46 - 0.72 | 0.54 [0.47 - 0.68] | [1] |
| theta_ini ($\theta_t$) | Rate of treatment initiation from symptomatic TB per year (initial) | 0.00 - 0.57 | 0.46 [0.34 - 0.56] | - |
| theta_fin ($\theta_t$) | Rate of treatment initiation from symptomatic TB per year (final) | 0.57 - 0.77 | 0.71 [0.60 - 0.76] | [8] |
| delta (δ) | Rate of treatment completion per year | 2.00 | - | [16] |
| phi_ini ($\phi_t$) | Rate of treatment failure per year (initial) | 0.11 - 1.00 | 0.63 [0.21 - 0.97] | - |
| phi_fin ($\phi_t$) | Rate of treatment failure per year (final) | 0.07 - 0.11 | 0.09 [0.07 - 0.11] | [8] |

| | | | | |
|---|---|---|---|---|
| mutb_ini ($\mu_{TB,t}$) | TB-specific mortality rate per year (initial) | 0.28 - 0.38 | 0.34 [0.29 - 0.37] | [10] |
| mutb_fin ($\mu_{TB,t}$) | TB-specific mortality rate per year (final) | 0.00 - 0.28 | 0.17 [0.07 - 0.27] | - |
| mu ($\mu$) | Background mortality rate per year | 0.014 | - | - |

Model parameters description for deterministic TB transmission model calibrated to TB epidemiology in Viet Nam. Ranges for priors and median value with corresponding 95% uncertainty intervals (95%UI) for non-implausible ranges (posteriors) are shown. When range is not shown, constant value was used.

All parameters are expressed per year.

**Table S3. Probability of a positive test per model state for each screening tool.**

| Test | State | Value (Range) | Description |
|---|---|---|---|
| Nucleic acid amplification test (NAAT, Xpert MTB/RIF Ultra) | Susceptible | 0.006 (0.005 – 0.008) | (1 – specificity) for individuals in a community in Kampala, Uganda [17] |
| | Infected | 0.006 (0.005 – 0.008) | (1 – specificity) for individuals in a community in Kampala, Uganda [17] |
| | Cleared | 0.006 (0.005 – 0.008) | (1 – specificity) for individuals in a community in Kampala, Uganda [17] |
| | Recovered | 0.040 (0.020 – 0.060) | (1 – specificity) for individuals screened positive for symptoms and/or CXR with a history of TB in the community [18] |
| | Unconfirmed TB | 0.044 (0.026 – 0.070) | (1 – specificity) for pulmonary TB from individuals in primary care facilities and local hospitals [19] |
| | Asymptomatic TB | 0.775 (0.676 – 0.856) | Sensitivity for smear-negative TB from individuals in primary care facilities and local hospitals [19] |
| | Symptomatic TB | 0.909 (0.862 – 0.947) | Sensitivity for pulmonary TB from individuals in primary care facilities and local hospitals [19] |
| | Treated | 0.040 (0.020 – 0.060) | (1 – specificity) for individuals screened positive for symptoms and/or CXR with a history of TB in the community [18] |
| Chest radiography with CAD software interpretation (CXR) | Susceptible | 0.085 (0.069 – 0.134) | Median and interquartile range for the proportion of national TB prevalence survey participants with abnormal CXR, regardless of TB status [20] |
| | Infected | 0.085 (0.069 – 0.134) | Median and interquartile range for the proportion of national TB prevalence survey participants with abnormal CXR, regardless of TB status [20] |
| | Cleared | 0.085 (0.069 – 0.134) | Median and interquartile range for the proportion of national TB prevalence survey participants with abnormal CXR, regardless of TB status [20] |
| | Recovered | 0.503 (0.481 – 0.524) | Proportion with abnormal CXR suggestive of TB among participants of national TB prevalence survey reporting TB history [21] |
| | Unconfirmed TB | 0.677* (0.626 – 0.712) | Midpoint between the median and bounds of values for Recovered/Treated and Symptomatic TB [Assumption] |
| | Asymptomatic TB | 0.677* (0.626 – 0.712) | Midpoint between the median and bounds of values for Recovered/Treated and Symptomatic TB [Assumption] |
| | Symptomatic TB | 0.910 (0.900 – 0.920) | Sensitivity of CAD software for bacteriologically confirmed TB in screening use case [22] |
| | Treated | 0.503 (0.481 – 0.524) | Proportion with abnormal CXR suggestive of TB among participants of national TB prevalence survey reporting TB history [21] |

| | | | |
|---|---|---|---|
| Nucleic acid amplification test (NAAT, Xpert MTB/RIF) | Susceptible | 0.0022 (0.0016 – 0.0029) | (1 – specificity) for individuals in a community in selected villages in Ca Mau province, Viet Nam [23] |
| | Infected | 0.0022 (0.0016 – 0.0029) | (1 – specificity) for individuals in a community in selected villages in Ca Mau province, Viet Nam [23] |
| | Cleared | 0.0022 (0.0016 – 0.0029) | (1 – specificity) for individuals in a community in selected villages in Ca Mau province, Viet Nam [23] |
| | Recovered | 0.026 (0.005 – 0.083) | (1 – specificity) for individuals with a history of TB in primary care facilities and local hospitals [19] |
| | Unconfirmed TB | 0.016 (0.007 – 0.030) | (1 – specificity) for pulmonary TB from individuals in primary care facilities and local hospitals [19] |
| | Asymptomatic TB | 0.606 (0.484 – 0.717) | Sensitivity for smear-negative TB from individuals in primary care facilities and local hospitals [19] |
| | Symptomatic TB | 0.847 (0.786 – 0.899) | Sensitivity for pulmonary TB from individuals in primary care facilities and local hospitals [19] |
| | Treated | 0.026 (0.005 – 0.083) | (1 – specificity) for individuals with a history of TB in primary care facilities and local hospitals [19] |
| Further investigation (informed by prolonged cough) | Susceptible | 0.061 (0.047 – 0.074) | Median and interquartile range for the proportion of national TB prevalence survey participants reporting prolonged cough, regardless of CXR or TB status [20] |
| | Infected | 0.061 (0.047 – 0.074) | Median and interquartile range for the proportion of national TB prevalence survey participants reporting prolonged cough, regardless of CXR or TB status [20] |
| | Cleared | 0.061 (0.047 – 0.074) | Median and interquartile range for the proportion of national TB prevalence survey participants reporting prolonged cough, regardless of CXR or TB status [20] |
| | Recovered | 0.131 (0.089 – 0.162) | Midpoint between the median and bounds of values for S/I/C and Unconfirmed TB [Assumption] |
| | Unconfirmed TB | 0.201 (0.129 – 0.249) | Median and interquartile range for the proportion of national TB prevalence survey participants reporting prolonged cough with abnormal CXR, regardless of TB status [20] |
| | Asymptomatic TB | 1.00 | Assuming a strong clinical appraisal that is able to diagnose confirmed TB [Assumption] |
| | Symptomatic TB | 1.00 | Assuming a strong clinical appraisal that is able to diagnose confirmed TB [Assumption] |
| | Treated | 0.566 (0.545 – 0.581) | Midpoint between the median and bounds of values for Recovered and Asymptomatic/Symptomatic TB [Assumption] |

Probability of a positive test result for each screening diagnostic tool for each state in the model and were independently sampled from uniform distributions for each model run. Test positivity under further investigation refers to individuals who have tested positive based on a given screening algorithm. *Under a sensitivity analysis, evaluating revised CXR sensitivity for *Unconfirmed* and *Asymptomatic* TB, the value matches the one in *Recovered* and *Treated*. CAD: Computer-aided diagnosis; TB: Tuberculosis.

**Table S4. Costing estimates for business-as-usual TB diagnosis and treatment.**

| Cost type | Cost per individual (US$) | Distribution | Notes |
|---|---|---|---|
| Diagnosis for DS-TB | 264.0 | Gamma distribution, standard deviation 20% of the mean | Considering NNT, CXR, bacteriological costs, and staff time costs |
| Diagnosis for DR-TB | 1595.0 | Gamma distribution, standard deviation 20% of the mean | Considering NNT, CXR, bacteriological costs, and staff time costs |
| Treatment for DS-TB | 81.0 | Gamma distribution, standard deviation 20% of the mean | Includes TB drugs, healthcare staff, bacterial monitoring, and overhead costs per treatment episode |
| Treatment for DR-TB | 973.0 | Gamma distribution, standard deviation 20% of the mean | Includes TB drugs, healthcare staff, bacterial monitoring, and overhead costs per treatment episode |

Costing estimates per individual for business-as-usual TB diagnosis and treatment provided by the national TB programme in Viet Nam. Costs were independently sampled from gamma distributions as specified for each model run. CXR: Chest radiography; DR-TB: Drug-resistant TB; DS-TB: Drug-susceptible TB; NNT: Number needed to test; TB: Tuberculosis; US$: United States dollar.

**Table S5. Costing estimates for population-wide screening algorithms.**

| Analysis | Algorithm | Cost per individual (US$) | Distribution |
|---|---|---|---|
| Main analysis | NAAT-only | 8.0 | Gamma distribution, standard deviation 20% of the mean |
| | CXR+NAAT | 1.7 | Gamma distribution, standard deviation 20% of the mean |
| | CXR-only | 1.2 | Gamma distribution, standard deviation 20% of the mean |
| Sensitivity analysis | NAAT-only (US$1 NAAT) | 3.0 | Gamma distribution, standard deviation 20% of the mean |
| | CXR+NAAT (US$1 NAAT) | 1.3 | Gamma distribution, standard deviation 20% of the mean |

Costing estimates per individual for different population-wide algorithms. Estimates represent the average cost per individual screened, calculated from the total costs of six years of community-wide screening interventions, informed by the ACT3 trial and the ongoing ACT5 trial [11,24]. Costs account for several factors, including the number of screening days per year, human resource costs (e.g., technicians, field workers, laboratory staff, administrative staff, and supervisors), the proportion of the population participating, the proportion providing sputum samples, the number undergoing NAAT, consumables, setup of screening sites, and transportation. For the CXR+NAAT algorithm, costs also include the proportion of CXR deemed abnormal, requiring confirmatory NAAT testing. Unit costs were independently sampled from a gamma distribution. NAAT: Nucleic acid amplification test; CXR: Chest radiography; US$: United States dollar.

**Table S6. Performance of population-wide screening interventions to reach TB prevalence threshold of 100 per 100,000 inhabitants.**

| Screening algorithm | BAU | NAAT | | NAAT+CXR | | CXR |
|---|---|---|---|---|---|---|
| **Rounds required to reach threshold** | Not reached | 3 annual rounds | | 3 annual rounds | | 2 annual rounds |
| **Cumulative TB incidence** | 2.25m (95%UI: 1.57-3.04) | 1.39m (95%UI: 0.94-1.89) | | 1.52m (95%UI: 1.04-2.06) | | 0.79m (95%UI: 0.55-1.10) |
| **Cumulative TB deaths** | 273k (95%UI:123-475) | 160k (95%UI: 70-278) | | 177k (95%UI: 77-308) | | 94k (95%UI:41-163) |
| **Cumulative DALYs** | 8.12m (95%UI: 5.85-10.83) | 5.13m (95%UI: 3.60-6.82) | | 5.62m (95%UI: 4.04-7.37) | | 3.09m (95%UI: 2.21-4.17) |
| **Cumulative TPs diagnosed through screening** | N/A | 369k (95%UI: 281-450) | | 313k (95%UI: 232-392) | | 988k (95%UI: 666-1,307) |
| **Cumulative FPs diagnosed through screening** | N/A | 1,384k (95%UI: 1,033-1,834) | | 617k (95%UI: 360-983) | | 21,107k (95%UI: 16,340-25,722) |
| **Unit price of NAAT** | N/A | US$8 | US$1 | US$8 | US$1 | N/A |
| **Cost of diagnosis/screening** | 363m (95%UI: 222-578) | 1,343m (95%UI: 952-1,846) | 641m (95%UI: 471-842) | 639m (95%UI: 478-858) | 548m (95%UI: 398-719) | 311m (95%UI: 230-411) |
| **Cost of treatment** | 138m (95%UI: 86-209) | 235m (95%UI: 157-344) | | 176m (95%UI: 114-268) | | 1,806m (95%UI: 1,133-2,796) |
| **Budget impact** | 505m (95%UI: 328-757) | 1,583m (95%UI: 1,183-2,102) | 878m (95%UI: 677-1,113) | 822m (95%UI: 617-1,075) | 722m (95%UI: 548-932) | 2,118m (95%UI: 1,451-3,093) |
| **Annual cost of front-loading** | N/A | 429m (95%UI: 297-586) | 192m (95%UI: 139-255) | 162m (95%UI: 113-219) | 131m (95%UI: 93-183) | 967m (95%UI: 646-1,466) |
| **Annual cost savings** | N/A | 8.0m (95%UI: 1.9-15.8) | | 6.9m (95%UI: 0.3-14.8) | | 13.0m (95%UI: 7.1-21.8) |
| **ICER compared with BAU (US$ per DALY averted)** | N/A | 354 (95%UI: 144-811) | 123 (95%UI: 24-325) | 123 (95%UI: 21-359) | 84 (95%UI: 1-285) | 318 (95%UI: 133-724) |

Epidemiological performance and economic impact of population-wide screening interventions in Viet Nam per algorithm when conducted to reach TB prevalence threshold of 100 per 100,000 people. The values represent cumulative accrual over the 25-year time horizon, extending up to 2050. Budget impact includes cumulative costs of screening/diagnosis and treatment for both BAU and the intervention algorithm up to 2050. The cost of front-loading refers to intervention-specific screening and treatment costs, presented as the annual average during the intervention period. Annual cost savings are defined as the difference in BAU-specific diagnosis and treatment costs between the intervention algorithm and the BAU counterfactual, averaged over the time horizon. BAU: Business-as-usual; CXR: Chest radiography; DALY: Disability-adjusted life year; FP: False positive; ICER: Incremental cost-effectiveness ratio; NAAT: Nucleic acid amplification test (Xpert MTB/RIF Ultra); TB: Tuberculosis; TP: True positive; UI: Uncertainty interval; US$: United States dollar.

**Table S7. Performance of population-wide screening interventions to reach TB prevalence threshold of 20 per 100,000 inhabitants.**

| Screening algorithm | BAU | NAAT | | NAAT+CXR | | CXR |
|---|---|---|---|---|---|---|
| **Rounds required to reach threshold** | Not reached | 11 annual rounds | | 12 annual rounds | | 4 annual rounds |
| **Cumulative TB incidence** | 2.25m (95%UI: 1.57-3.04) | 0.62m (95%UI: 0.42-0.85) | | 0.68m (95%UI: 0.47-0.93) | | 0.36m (95%UI: 0.26-0.49) |
| **Cumulative TB deaths** | 273k (95%UI:123-475) | 63k (95%UI: 26-107) | | 68k (95%UI: 29-113) | | 43k (95%UI:20-74) |
| **Cumulative DALYs** | 8.12m (95%UI: 5.85-10.83) | 2.92m (95%UI: 2.06-3.84) | | 3.25m (95%UI: 2.35-4.31) | | 1.77m (95%UI: 1.30-2.35) |
| **Cumulative TPs diagnosed through screening** | N/A | 711k (95%UI: 520-907) | | 718k (95%UI: 504-946) | | 1,262k (95%UI: 852-1,676) |
| **Cumulative FPs diagnosed through screening** | N/A | 5,276k (95%UI: 3,917-6,834) | | 3,022k (95%UI: 1,754-4,667) | | 42,402k (95%UI: 32,130-51,051) |
| **Unit price of NAAT** | N/A | US$8 | US$1 | US$8 | US$1 | N/A |
| **Cost of diagnosis/screening** | 363m (95%UI: 222-578) | 4,259m (95%UI: 2,782-6,223) | 1,671m (95%UI: 1,097-2,380) | 2,031m (95%UI: 1,375-2,873) | 1,555m (95%UI: 1,061-2,243) | 420m (95%UI: 293-577) |
| **Cost of treatment** | 138m (95%UI: 86-209) | 540m (95%UI: 336-827) | | 349m (95%UI: 215-562) | | 3,494m (95%UI: 2,148-5,477) |
| **Budget impact** | 505m (95%UI: 328-757) | 4,801m (95%UI: 3,301-6,780) | 2,219m (95%UI:1,586-2,973) | 2,368m (95%UI: 1,692-3,259) | 1,929m (95%UI: 1,387-2,599) | 3,913m (95%UI: 2,598-5,873) |
| **Annual cost of front-loading** | N/A | 426m (95%UI: 290-605) | 191m (95%UI: 136-259) | 187m (95%UI: 131-260) | 150m (95%UI: 106-207) | 958m (95%UI: 634-1,447) |
| **Annual cost savings** | N/A | 15.5m (95%UI: 9.0-24.9) | | 15.2m (95%UI: 8.8-24.7) | | 16.9m (95%UI: 10.6-26.6) |
| **ICER compared with BAU (US$ per DALY averted)** | N/A | 825 (95%UI: 380-1,713) | 328 (95%UI: 143-688) | 381 (95%UI: 172-818) | 291 (95%UI: 122-643) | 537 (95%UI: 240-1,207) |

Epidemiological performance and economic impact of population-wide screening interventions in Viet Nam per algorithm when conducted to reach TB prevalence threshold of 20 per 100,000 people. The values represent cumulative accrual over the 25-year time horizon, extending up to 2050. Budget impact includes cumulative costs of screening/diagnosis and treatment for both BAU and the intervention algorithm up to 2050. The cost of front-loading refers to intervention-specific screening and treatment costs, presented as the annual average during the intervention period. Annual cost savings are defined as the difference in BAU-specific diagnosis and treatment costs between the intervention algorithm and the BAU counterfactual, averaged over the time horizon. BAU: Business-as-usual; CXR: Chest radiography; DALY: Disability-adjusted life year; FP: False positive; ICER: Incremental cost-effectiveness ratio; NAAT: Nucleic acid amplification test (Xpert MTB/RIF Ultra); TB: Tuberculosis; TP: True positive; UI: Uncertainty interval; US$: United States dollar.

**Table S8. Performance of population-wide screening interventions with further investigation post-screening.**

| Screening algorithm | BAU | NAAT | CXR+NAAT | CXR |
|---|---|---|---|---|
| **Rounds required to reach threshold** | Not reached | 6 annual rounds | 8 annual rounds | 3 annual rounds |
| **Cumulative TB incidence** | 2.25m (95%UI: 1.57-3.04) | 1.03m (95%UI: 0.68-1.41) | 1.04m (95%UI: 0.69-1.43) | 1.22m (95%UI: 0.83-1.67) |
| **Cumulative TB deaths** | 273k (95%UI:123-475) | 113k (95%UI: 47-194) | 112k (95%UI: 46-196) | 140k (95%UI: 62-248) |
| **Cumulative DALYs** | 8.12m (95%UI: 5.85-10.83) | 3.99m (95%UI: 2.77-5.31) | 4.17m (95%UI: 2.89-5.56) | 4.52m (95%UI: 3.22-6.06) |
| **Cumulative TPs diagnosed through screening** | N/A | 490k (95%UI: 377-594) | 489k (95%UI: 364-600) | 514k (95%UI: 374-670) |
| **Cumulative FPs diagnosed through screening** | N/A | 272k (95%UI: 182-410) | 224k (95%UI: 127-383) | 3,057k (95%UI: 2,236-4,051) |
| **Unit price of NAAT** | N/A | 8USD | 8USD | N/A |
| **Cost of diagnosis/screening** | 363m (95%UI: 222-578) | 2,399m (95%UI: 1,655-3,405) | 1,243m (95%UI: 816-1,746) | 478m (95%UI: 339-641) |
| **Cost of treatment** | 138m (95%UI: 86-209) | 137m (95%UI: 91-204) | 134m (95%UI: 84-203) | 369m (95%UI: 237-553) |
| **Budget impact** | 505m (95%UI: 328-757) | 2,540m (95%UI: 1,794-3,567) | 1,377m (95%UI: 955-1,885) | 921m (95%UI: 688-1,177) |
| **Annual cost of front-loading** | N/A | 387m (95%UI: 262-555) | 145m (95%UI: 95-209) | 193m (95%UI: 139-262) |
| **Annual cost savings** | N/A | 11.7m (95%UI: 5.8-20.3) | 11.6m (95%UI: 5.9-20.2) | 9.5m (95%UI: 3.8-18.0) |
| **ICER compared with BAU (US$ per DALY averted)** | N/A | 489 (95%UI: 207-1,105) | 219 (95%UI: 73-521) | 113 (95%UI: 28-278) |

Epidemiological performance and economic impact of population-wide screening interventions with further investigation post-screening in Viet Nam per algorithm when conducted to reach TB prevalence threshold of 50 per 100,000 people. Further investigation was applied and costed exclusively for individuals who screened positive under their respective algorithm. The values represent cumulative accrual over the 25-year time horizon, extending up to 2050. Budget impact includes cumulative costs of screening/diagnosis and treatment for both BAU and the intervention algorithm up to 2050. The cost of front-loading refers to intervention-specific screening and treatment costs, presented as the annual average during the intervention period. Annual cost savings are defined as the difference in BAU-specific diagnosis and treatment costs between the intervention algorithm and the BAU counterfactual, averaged over the time horizon. BAU: Business-as-usual; CXR: Chest radiography; DALY: Disability-adjusted life year; FP: False positive; ICER: Incremental cost-effectiveness ratio; NAAT: Nucleic acid amplification test (Xpert MTB/RIF Ultra); TB: Tuberculosis; TP: True positive; UI: Uncertainty interval; US$: United States dollar.

**Table S9. Performance of population-wide screening interventions with revised CXR sensitivity.**

| Screening algorithm | BAU | NAAT | CXR+NAAT | CXR |
|---|---|---|---|---|
| **Rounds required to reach threshold** | Not reached | 6 annual rounds | 9 annual rounds | 3 annual rounds |
| **Cumulative TB incidence** | 2.25m (95%UI: 1.57-3.04) | 0.95m (95%UI: 0.63-1.31) | 1.00m (95%UI: 0.67-1.35) | 0.68m (95%UI: 0.47-0.93) |
| **Cumulative TB deaths** | 273k (95%UI:123-475) | 104k (95%UI: 44-184) | 105k (95%UI: 45-185) | 79k (95%UI: 34-139) |
| **Cumulative DALYs** | 8.12m (95%UI: 5.85-10.83) | 3.74m (95%UI: 2.64-4.99) | 4.14m (95%UI: 2.91-5.52) | 2.79m (95%UI: 2.00-3.74) |
| **Cumulative TPs diagnosed through screening** | N/A | 555k (95%UI: 411-688) | 544k (95%UI: 395-698) | 985k (95%UI: 672-1,305) |
| **Cumulative FPs diagnosed through screening** | N/A | 2,779k (95%UI: 2,059-3,696) | 1,924k (95%UI: 1,122-3,013) | 31,354k (95%UI: 24,199-38,227) |
| **Unit price of NAAT** | N/A | US$8 | US$8 | N/A |
| **Cost of diagnosis/screening** | 363m (95%UI: 222-578) | 2,428m (95%UI: 1,675-3,465) | 1,350m (95%UI: 945-1,881) | 374m (95%UI: 273-509) |
| **Cost of treatment** | 138m (95%UI: 86-209) | 336m (95%UI: 220-511) | 272m (95%UI: 158-425) | 2,570m (95%UI: 1,623-4,103) |
| **Budget impact** | 505m (95%UI: 328-757) | 2,766m (95%UI: 1,965-3,782) | 1,639m (95%UI: 1,191-2,196) | 2,967m (95%UI: 2,005-4,487) |
| **Annual cost of front-loading** | N/A | 427m (95%UI: 299-599) | 159m (95%UI: 110-219) | 938m (95%UI: 623-1,448) |
| **Annual cost savings** | N/A | 12.3m (95%UI: 6.5-21.4) | 12.2m (95%UI: 5.9-21.0) | 14.3m (95%UI: 8.1-23.2) |
| **ICER compared with BAU (US$ per DALY averted)** | N/A | 516 (95%UI: 233-1,073) | 283 (95%UI: 114-640) | 456 (95%UI: 204-1,062) |

Epidemiological performance and economic impact of population-wide screening interventions with revised CXR sensitivity for unconfirmed and asymptomatic TB in Viet Nam per algorithm when conducted to reach TB prevalence threshold of 50 per 100,000 people. The values represent cumulative accrual over the 25-year time horizon, extending up to 2050. Budget impact includes cumulative costs of screening/diagnosis and treatment for both BAU and the intervention algorithm up to 2050. The cost of front-loading refers to intervention-specific screening and treatment costs, presented as the annual average during the intervention period. Annual cost savings are defined as the difference in BAU-specific diagnosis and treatment costs between the intervention algorithm and the BAU counterfactual, averaged over the time horizon. BAU: Business-as-usual; CXR: Chest radiography; DALY: Disability-adjusted life year; FP: False positive; ICER: Incremental cost-effectiveness ratio; NAAT: Nucleic acid amplification test (Xpert MTB/RIF Ultra); TB: Tuberculosis; TP: True positive; UI: Uncertainty interval; US$: United States dollar.

**Table S10. Cost-effectiveness of population-wide screening interventions for TB.**

| Analysis type | Screening algorithm | DALYs averted compared to BAU | Additional costs (US$) compared to BAU | Incremental DALYs | Incremental costs (US$) | ICER (US$ per DALY averted) |
|---|---|---|---|---|---|---|
| Reducing the unit price of NAAT cartridges to US$1 | CXR+NAAT | 4.28m (95%UI: 2.93-6.16) | 0.71b (95%UI: 0.35-1.11) | 4.28m (95%UI: 2.93-6.16) | 0.71b (95%UI: 0.35-1.11) | 167 (95%UI: 57-380) |
| | NAAT | 4.38m (95%UI: 2.97-6.19) | 0.83b (95%UI: 0.44-1.27) | Removed due to extended dominance with respect to CXR-only | | |
| | CXR | 5.94m (95%UI: 4.18-7.97) | 2.44b (95%UI: 1.41-3.88) | 1.52m (95%UI: 0.79-2.37) | 1.61b (95%UI: 0.51-3.06) | 1,057 (95%UI: 642-1,291) |
| TB prevalence threshold of 100 per 100,000 people | CXR+NAAT | 2.55m (95%UI: 1.53-3.92) | 0.31b (95%UI: 0.08-0.55) | 2.55m (95%UI: 1.53-3.92) | 0.31b (95%UI: 0.08-0.55) | 123 (95%UI: 21-359) |
| | NAAT | 3.02m (95%UI: 1.93-4.47) | 1.07b (95%UI: 0.64-1.57) | Removed due to extended dominance with respect to CXR-only | | |
| | CXR | 5.06m (95%UI: 3.56-6.86) | 1.61b (95%UI: 0.91-2.58) | 2.49m (95%UI: 1.57-3.54) | 1.31b (95%UI: 0.59-2.25) | 528 (95%UI: 376-636) |
| TB prevalence threshold of 20 per 100,000 people | CXR+NAAT | 4.89m (95%UI: 3.34-6.92) | 1.86b (95%UI: 1.19-2.73) | 4.89m (95%UI: 3.34-6.92) | 1.86b (95%UI: 1.19-2.73) | 381 (95%UI: 172-818) |
| | NAAT | 5.19m (95%UI: 3.65-7.35) | 4.29b (95%UI: 2.79-6.26) | Removed due to simple dominance with respect to CXR-only | | |
| | CXR | 6.32m (95%UI: 4.49-8.66) | 3.39b (95%UI: 2.08-5.43) | 1.46m (95%UI: 0.81-2.31) | 1.52b (95%UI: 0.12-3.62) | 1,036 (95%UI: 148-1,567) |
| Performance of using Xpert MTB/RIF in a NAAT-only algorithm | NAAT (Xpert MTB/RIF) | 4.29m (95%UI: 2.93-6.16) | 2.96b (95%UI: 1.89-4.41) | Removed due to simple dominance with respect to CXR+NAAT | | |
| | CXR+NAAT | 4.29m (95%UI: 2.86-6.14) | 0.97b (95%UI: 0.52-1.49) | 4.29m (95%UI: 2.86-6.14) | 0.97b (95%UI: 0.52-1.49) | 225 (95%UI: 85-520) |
| | NAAT (Xpert MTB/RIF Ultra) | 4.36m (95%UI: 3.09-6.23) | 2.25b (95%UI: 1.45-3.31) | Removed due to extended dominance with respect to CXR-only | | |
| | CXR | 5.94m (95%UI: 4.18-7.97) | 2.44b (95%UI: 1.41-3.88) | 1.61m (95%UI: 0.86-2.56) | 1.49b (95%UI: 0.34-2.87) | 927 (95%UI: 393-1,124) |

| | | | | | | |
|---|---|---|---|---|---|---|
| Performance of further investigation post-screening | CXR | 3.59m (95%UI: 2.43-5.23) | 0.41b (95%UI: 0.15-0.68) | 3.59m (95%UI: 2.43-5.23) | 0.41b (95%UI: 0.15-0.68) | 113 (95%UI: 28-278) |
| | CXR+NAAT | 3.96m (95%UI: 2.61-5.83) | 0.87b (95%UI: 0.43-1.36) | 0.37m (95%UI: 0.00-1.15) | 0.46b (95%UI: 0.03-1.00) | 1,293 (95%UI: 1,153-1,555) |
| | NAAT | 4.14m (95%UI: 2.77-5.99) | 2.03b (95%UI: 1.24-3.06) | 0.16m (95%UI: 0.00-0.48) | 1.16b (95%UI: 0.23-2.23) | 6,183 (95%UI: 5,165-10,441) |
| Revised CXR sensitivity for unconfirmed and asymptomatic TB | CXR+NAAT | 3.98m (95%UI: 2.64-5.73) | 1.12b (95%UI: 0.65-1.69) | 3.98m (95%UI: 2.64-5.73) | 1.12b (95%UI: 0.65-1.69) | 283 (95%UI: 113-640) |
| | NAAT | 4.36m (95%UI: 3.09-6.23) | 2.25b (95%UI: 1.45-3.31) | Removed due to extended dominance with respect to CXR-only | | |
| | CXR | 5.35m (95%UI: 3.77-7.33) | 2.44b (95%UI: 1.49-4.00) | 1.34m (95%UI: 0.62-2.16) | 1.32b (95%UI: 0.22-2.95) | 962 (95%UI: 750-1,447) |

Cost-effectiveness sensitivity analyses of population-wide screening interventions in Viet Nam per algorithm. The values represent cumulative accrual over the 25-year time horizon, extending up to 2050. BAU: Business-as-usual; CXR: Chest radiography with computer-aided detection software interpretation; DALY: Disability-adjusted life year; ICER: Incremental cost-effectiveness ratio; NAAT: Nucleic acid amplification test (primarily Xpert MTB/RIF Ultra unless specified otherwise); TB: Tuberculosis; UI: Uncertainty interval; US$: United States dollar.

15. Verver S, Warren RM, Beyers N, Richardson M, van der Spuy GD, Borgdorff MW, et al. Rate of reinfection tuberculosis after successful treatment is higher than rate of new tuberculosis. Am J

Respir Crit Care Med. 2005;171: 1430–1435. doi:10.1164/rccm.200409-1200OC

16. World Health Organization. WHO Consolidated Guidelines on Tuberculosis. Module 4: Treatment - Drug-susceptible tuberculosis treatment. Geneva, Switzerland: WHO; 2022.

22. World Health Organization. WHO Consolidated Guidelines on Tuberculosis. Module 2: Screening - Systematic screening for tuberculosis disease. Geneva, Switzerland: WHO; 2021 Mar.

23. Ho J, Nguyen PTB, Nguyen TA, Tran KH, Van Nguyen S, Nguyen NV, et al. Reassessment of the positive predictive value and specificity of Xpert MTB/RIF: a diagnostic accuracy study in the context of community-wide screening for tuberculosis. Lancet Infect Dis. 2016;16: 1045–1051. doi:10.1016/S1473-3099(16)30067-6

24. Australian New Zealand Clinical Trials Registry. ACTRN12622000115730. In: ANZCTR [Internet]. [cited 6 Feb 2024]. Available: https://anzctr.org.au/Trial/Registration/TrialReview.aspx?ACTRN=12622000115730
